# Assessment of pediatric surgical needs, health seeking behaviors and health systems in the rural district of Tando Mohammad Khan Sindh, Pakistan

**DOI:** 10.1101/2022.06.28.22277027

**Authors:** Saqib Hamid Qazi, Syed Saqlain Ali Meerza, Reinou S. Groen, Sohail Asghar Dogar, Mushtaq Mirani, Muhammad Khan Jamali, Zahid Ali Khan, Zahra Ali Padhani, Rasool Bux, Imran Ahmed Chahudary, Arjumand Rizvi, Saleem Islam, Sadaf Khan, Rizwan Haroon Ur Rashid, Syed Akbar Abbas, Abdul Sami Memon, Sadia Tabassum, Bukhtawar Dilawar, Jai K Das

**Affiliations:** Section of Pediatric Surgery, Department of Surgery, Aga Khan University, Karachi, Pakistan; Institute for Global Health and Development, Aga Khan University, Karachi, Pakistan; Department of Obstetrics and Gynecology, Alaska Native Medical Center, Anchorage, Alaska; Division of Women and Child Health, Aga Khan University Hospital, Karachi, Pakistan; Department of Pediatric Surgery, University of Florida, Gainesville, United States of America; Department of Surgery, Aga Khan University, Karachi, Pakistan; Section of Orthopedic Surgery, Department of Surgery, Aga Khan University, Karachi, Pakistan; Section of Head and Neck Surgery, Department of Surgery Aga Khan University, Karachi, Pakistan; Department of Ophthalmology and Visual Sciences, Aga Khan University, Karachi, Pakistan; Section of Dermatology, Department of Medicine, Aga Khan University, Karachi, Pakistan

**Author notes:** **Address correspondence to:** Dr Jai K. Das, Assistant Director, Institute for Global Health and Development, Aga Khan University, Karachi, Pakistan.

## Abstract

**Background:** Surgical conditions are responsible for up to 15% of total DALY lost globally. Worldwide estimates have found that approximately 4.8 billion people have no access to surgical care. Within South Asia, greater than 95% of the population does not have access to care for conditions that require surgical management. Considering that greater than 50% of the population in the least developed regions worldwide is children, the surgical burden amongst children in LMICs is immense. In this study we use the SOSAS and PediPIPES in TMK district to assess the surgical needs of children under-5, quality of health facilities, and care seeking behavior in the community.

**Material and Methods:** The research was reviewed and approved by the Aga Khan University (AKU) Ethical Review Committee (ERC) and the National Bioethics Committee (NBC). Confidentiality of all collected data was assigned high priority at each stage of data handling. Data was collected through the SOSAS and PediPIPES survey tools between November 2019 and February 2020 from a total of 3,643 households in the TMK, Sindh, Pakistan. The SOSAS survey was conducted by research associates trained for data collection. Household mothers provided information about their children and data was recorded electronically. Health facilities were assessed using PediPIPES survey form. Information was collected on hard copies from all 39 health care facilities in the district, including RHCs, BHUs, DCDs, and DHQ. Data was collected by core team and entered onto an excel sheet.

**Results:** A total of 3,643 households participated and information of 6,371 children was collected. A total of 1,794 children were identified to have 3,072 lesions that required surgical attention. We categorized the lesions requiring surgical care according to six regions of the body. Head and neck accounted for the greatest number of lesions (n = 1,697) and the most significant unmet surgical need (16.6%). The chest region had 102 lesions and the least unmet surgical need of 5.9%. The back accounted for 87 lesions with an unmet surgical need of 6.9%. The abdomen had 493 lesions and an unmet surgical need of 13.4%. A total of 169 lesions were found on buttocks/groin/genitalia region with an unmet surgical need of 14.8%, while extremities presented with 296 lesions amounting to 11.8% unmet surgical need.

A total of 39 health facilities, consisting of one DHQ, three RHCs, 14 BHUs and 21 DCDs, were surveyed. Trained staff were only present at the DHQ. Basic procedures such as suturing, wound debridement, I&D were performed more commonly than the more complex procedures. Most hospitals were found to have a good availability of equipment and supplies. PediPIPES scores and indices were calculated for the 39 health facilities in the area. The DHQ was found to have the highest score.

**Conclusions:** This study holds great significance for evaluation of pediatric surgical burden in Pakistan. It provides important insight into the burden of children’s operative disease in Pakistan’s rural district of TMK. The results show a significant need for provision of surgical care and has important implications for the global operative community as well as for strengthening the local health system in Pakistan. This data is useful preliminary evidence that emphasizes the need to further evaluate interventions for strengthening surgical systems in rural Pakistan.

## INTRODUCTION

Surgical conditions are responsible for up to 15% of total disability adjusted life years (DALYs) lost globally (1). Worldwide estimates have found that approximately 4.8 billion people have no access to surgical care, and within South Asia, greater than 95% of the population does not have access to care for conditions that require surgical management (2). Considering that greater than 50% of the population in the least developed regions worldwide is children, we can surmise that the surgical burden amongst children in LMICs is immense (3, 4). Currently, a large disproportion exists between the wealthiest and poorest third of the population globally, with the wealthy receiving a major share of 73.6% of surgical procedures and the poor receiving only 3.5% (5). Within poor countries, surgical services are concentrated almost wholly in cities and reserved largely for those who can pay for them (1). Until recently, pediatric surgical care in low and middle-income countries (LMICs) was largely overlooked, with global health attention primarily addressing communicable diseases, and maternal and infant mortality (5). However, improvement of surgical care delivery for children is now being prioritized as a fundamental component of health care in LMICs (6). Improving surgical care delivery also has significant economic and welfare benefits for the population, as untreated surgical conditions increase medical costs, disability, and death (4). Hence, development of methods to enhance the quality of pediatric surgical and trauma care in low-resource regions can remarkably decrease childhood morbidity and mortality (7) and alleviate the associated financial and emotional stress.

To promote the development of LMIC health care delivery systems that includes pediatric surgical care, it is vital to understand the local burden of surgical disease in children, individual understanding of these disease pathologies, and the capacity of health care facilities to handle the burden of disease. It is crucial to understand the pediatric surgical health related behaviors of the community, as children lack the ability to express their health issues, and their health-related decisions are mostly handled by parents and families. Surveys completed at the community level provide a thorough measure of real-time surgical need (4). The Surgeons OverSeas Assessment of Surgical Need (SOSAS) survey has been the most frequently used population-based survey to estimate the surgical disease burden. It has previously been used in countries like Uganda, Rwanda, Nepal, and Sierra Leone (8-15). The Pediatric Personnel, Infrastructure, Procedure, Equipment, and Supplies (PediPIPES) survey tool is designed to assess pediatric surgical capacity. This survey originated as the World Health Organization (WHO) tool for situational analysis but was subsequently redesigned by Surgeons OverSeas (SOS) to include absolute numbers of hospital beds and operating rooms, a binary system of measurement to allow easier counting of items, removing reasons for not performing procedures, and restructuring of individual questions (16).

Pakistan, a South Asian country with a population of approximately 221 million people is an LMIC with a Gross National Income GNI per capita of $1270 (17). Approximately 63% of the population lives in rural areas (18). Several studies have been published describing childhood surgical emergencies in Pakistan. However, through a literature review, it was identified that no community-based survey tool has yet been employed to determine Pakistan’s pediatric surgical disease burden in rural areas (19-21).

To better understand the ground reality of pediatric surgical disease burden in a low income region, keeping in mind the global trends of surgical care delivery depicted in past research, the objective of this study was to assess the burden of pediatric surgical disease, understand the local population’s health seeking behavior, and analyze the health care set-up in Tando Mohammad Khan, a rural district in the Sindh province of Pakistan. Using the SOSAS and PediPIPES survey form, our results would serve as preliminary evidence towards the requirement to further strengthen pediatric surgical health care delivery in Pakistan and would lay the foundation for targeted approaches towards improving pediatric surgical health education and access to surgical care facilities.

## METHODOLOGY

The survey was conducted in Tando Mohammad Khan district, Sindh over a period of three months, between 22^nd^ November 2019 and 28^th^ February 2020.

### Ethics Statement

The research was reviewed and approved by the Aga Khan University (AKU) Ethical Review Committee (ERC) and the National Bioethics Committee (NBC). Confidentiality of all collected data was assigned high priority at each stage of data handling. The research participants were informed about the purpose, methods and benefits and intended uses of the research. Informed verbal consent was obtained. Respondents were free to stop interviews at any time or skip any questions they did not want to answer. They had the right to ask questions at any point before, during or after the interview. All interviews were conducted by trained staff and in conditions of privacy. The respondents were informed about their rights. All data files were password-protected.

### Location

Sindh is the second largest province by population (approximately six million people) after Punjab, and the third largest province of Pakistan by total area (22). Tando Mohammad Khan is one of the 29 districts in Sindh with an area of 1,814 square kilometers (km^2^), overall population of 677,228 and a population density of 373 people/km^2^ (23)

### Survey Tools

The SOSAS survey consists of two portions. The first section collects demographic details from the head of the household, including age and sex of household members, and number of deaths in the household within the past year. Household members are those living in the same physical structure. The second half of the survey gathers information from mothers on both current and previous surgical conditions categorized into six distinct anatomical regions: face, head, and neck; chest and breast; abdomen; groin, genitals, and buttocks; back; and arms, hands, legs, and feet. Mothers answer questions based on whether they perceive their children as ever having had a surgical condition in at least one of these anatomic regions. Additional questions cover the type of injury/accident, timing of the condition, and health seeking behavior, which includes the type of health care sought, type of health care received, and reasons why care was not accessed. The survey questions were translated into Sindhi, the primary language of Tando Mohammad Khan.

The PIPES survey assesses gaps in the availability of essential and emergency surgical care (EESC) at the district health facilities. The data items were divided into five sections: Personnel, Infrastructure, Procedures, Equipment and Supplies (PIPES). This PediPIPES survey focuses on the capacity of health facilities to provide EESC to infants and children and has 118 total data items, compared to the 256 of the WHO tool.

### Data collection

Households in Tando Mohammad Khan districts were line-listed and a total of 3,643 eligible houses were randomized. The SOSAS survey was conducted by research associates who were trained for data collection and were monitored by senior managers. Verbal consent was obtained from the parents, and the surveys were administered in Sindhi. Mothers provided the survey information about their children below the age of five years. The information was recorded electronically via an application developed by the District Monitoring Unity (DMU).

Sindh’s health facilities include primary and secondary healthcare facilities. A total of 1,782 primary facilities in the province include rural health clinics (RHC), basic health units (BHU), dispensaries, Mother and Child health centers (MCH centers) amongst others (24). There are a total of 90 secondary healthcare facilities in Sindh including District Headquarter (DHQ) hospitals, Tehsil Headquarter (THQ) hospitals and other specialized secondary hospitals (25).

The PIPES survey data from all 39 health care facilities in Tando Mohammad Khan district, which included Rural Health Centers (RHC), Basic Health Units (BHU), District Council Dispensaries (DCD), and District Headquarter (DHQ) hospitals was collected by core team members which included a research coordinator and research specialist. The data was collected on hard copies and then entered onto an excel sheet.

Additionally, a total of 233 photographs were taken of children with lesions. These were reviewed by the relevant surgeons at the Aga Khan University Hospital for scoring and establishment of preliminary diagnoses.

### Data Analysis

Current surgical need was defined as a self-reported surgical problem present at the time of the interview. Unmet surgical need was defined as a current surgical problem for which the respondent did not access care. Descriptive analysis was performed using STATA 16 (Stata Statistical Software: Release 16. College Station, TX: StataCorp LP).

## RESULTS

A total of 3,643 households were surveyed for children’s surgical needs in Tando Muhammad Khan (TMK), a rural district in Sindh, Pakistan. Most families in the area comprised of 2-3 people (84.8%). Of these households, 3,821 mothers filled the Surgeons Overseas Assessment of Surgical Need (SOSAS) survey form. Majority of the mothers’ age ranged between 20-40 years (90.3%). Interviews with mothers identified a sum of 6,071 children in the area, with majority in the age range 1-5 years (n = 4,944, 81.4%). A near equal male (n = 3,063, 50.5%) to female ratio (n = 3,008, 49.5%) was present.

### Burden

#### Deaths

According to our survey, 290 of the 3,643 households reported death of a child in the last five years. Of these, 278 (95.9%) reported one death, 11 (3.8%) reported two deaths and only 1 (0.3%) reported a total of three deaths in the household. Overall, 300 child deaths were recorded, with most occurring in the age range 1-12 months (79.3%), and a near equal occurrence in both genders [Male (56.9%) and Female (43.1%)]. A total of 109 (36.3%) deaths occurred due to surgical conditions, of which majority were reported to have occurred due to congenital deformity (n= 33, 30.3%), a sum of 26 (23.9%) deaths occurred due to maternal bleeding or illness during birth, and 22 (20.2%) experienced abdominal distension or pain. The remaining 191 children (63.7%) did not report an apparent surgical disease process as the cause of death. In 5 children (4.6%) death resulted from an injury or an accident, which included animal attacks or falls.

The majority of these children (n= 87, 79.8%) were taken to a health care facility [Private facility (67.8%) or Government facility (29.9%)] for treatment prior to death, while others received management at home (2.3%). Their treatment focused primarily on medical management (n = 84, 96.6%). Only two patients (2.3%) underwent a major procedure and one (1.1%) had a minor procedure.

Most deaths were reported at home (n = 53, 48.6%) followed by health care facility (n = 46, 42.2%). The most common reason identified for not taking the child to a health care facility was lack of time (child died before any arrangement could be made, n = 9, 40.9%). Other reasons included perception of the conditions to be non-surgical (n = 4, 18.2 %) and no money for health facility (n = 3, 13.6%). **(Table 1)**

**Table 1:**
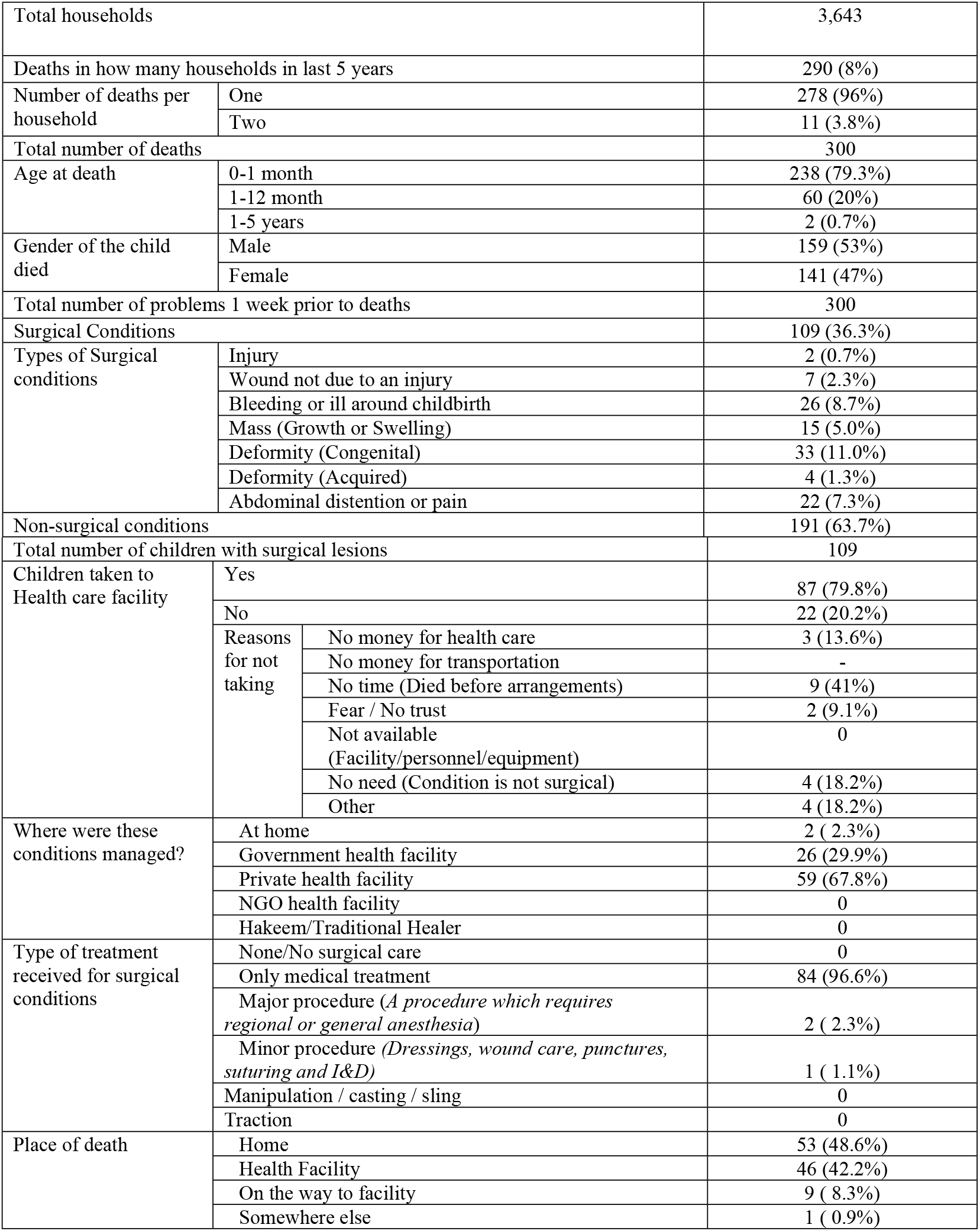
Details about child deaths in the district.

#### Lesions

Of the 6,071 children included in the survey, a total of 1,794 (29.6%) children were identified to have or previously have had a total number of 3,072 lesions requiring surgical care, as some children were identified to have more than one type of lesion. Lesions were approximately equally distributed among males (n = 1,615, 52.6%) and females (n = 1,457, 47.4%). Majority of the children affected were in the age range of 1-5 years (90.6%). An overview of the lesions is presented in **Table 2**. Through a head-to-toe inquiry via the SOSAS survey form, head and neck region (Comprising of head, eye, ear, and neck) was found to be most commonly affected (n =1,697, 55.2%) followed by extremities (n = 524, 17.1%). By far, the most common lesions identified overall were non-injury related wounds (n = 1,154, 37.6%) followed by mass/growth (n = 417, 13.6%). In the majority of cases (n = 2,538, 82.5%), the lesions were not secondary to an accident. Amongst the remaining 17.5% children who suffered an accident, falls were found to be the most common cause of injury (n = 264, 49.1%) followed by hot object/ hot liquid related injury (n = 126, 23.4%).

**Table 2:** Details of children with lesions.

|  |  |  |
| --- | --- | --- |
| Total number of children |  | 6071 |
| Total number of children with surgical lesions |  | 1794 (29.6%) |
| Total number of surgical lesions |  | 3072 |
| Age | 0-1 month | 5 (0.3%) |
|  | 1-12 months | 192 (10.7%) |
|  | 1-5 years | 1,597 (89.0%) |
| Gender of the child | Male | 939 (52.3%) |
|  | Female | 855 (47.7%) |

#### Head and neck

A total of 1,697 lesions identified in this region. Of these 697 (41.1%) were present on the ears, 383 (22.6%) on the head, 297 (17.5%) on the face, 204 (12%) in the eyes, and 116 (6.8%) lesions were on the neck. Lesions of the ear and face were mostly non-injury related, while the lesions on the head and the neck were primarily a mass or growth. Congenital deformity was the most common lesion of the eye. The majority of the head and neck lesions were not secondary to an accident or an injury (n = 1,526, 90%). In the remaining 171 (10%) accident-related injuries, fall (n = 106, 62%) was identified as the most common cause. The unmet surgical need was found to be the highest in the head and neck region, contributing to 16.6% of the total lesions. **(Table 3a)**

**Table 3a:** Lesions of head and neck.

| Region | Eyes |  | Ears |  | Face |  | Head |  | Neck |  |
| --- | --- | --- | --- | --- | --- | --- | --- | --- | --- | --- |
| What Problem he/she had | Children | Lesions | Children | Lesions | Children | Lesions | Children | Lesions | Children | Lesions |
| Wound injury related | 16 (8.2%) | 18 (8.8%) | 19 (3.6%) | 20 (2.9%) | 61 (22.1%) | 61 (20.5%) | 57 (16.1%) | 60 (15.7%) | 7 (6.3%) | 7 (6.0%) |
| Wound not injury related | 64 (32.7%) | 69 (33.8%) | 453 (86.8%) | 616 (88.4%) | 98 (35.5%) | 111 (37.4%) | 123 (34.8%) | 135 (35.2%) | 42 (37.5%) | 42 (36.2%) |
| Burn | 2 (1.0%) | 2 (1.0%) | 0(0) | 0(0) | 14 (5.1%) | 14 (4.7%) | 2 (0.6%) | 2 (0.5%) | 2 (1.8%) | 2 (1.7%) |
| Mass or growth (solid) | 11 (5.6%) | 12 (5.9%) | 21 (4.0%) | 26 (3.7%) | 81 (29.3%) | 88 (29.6%) | 146 (41.4%) | 160 (41.8%) | 49 (43.8%) | 53 (45.7%) |
| Deformity congenital | 80 (40.8%) | 80 (39.2%) | 20 (3.8%) | 25 (3.6%) | 11 (4.0%) | 11 (3.7%) | 13 (3.7%) | 13 (3.4%) | 5 (4.5%) | 5 (4.3%) |
| Deformity acquired | 10 (5.1%) | 10 (4.9%) | 6 (1.1%) | 6 (0.9%) | 11 (4.0%) | 12 (4.0%) | 12 (3.4%) | 13 (3.4%) | 7 (6.3%) | 7 (6.0%) |
| Blindness | 1 (0.5%) | 1 (0.5%) | 0(0) | 0(0) | 0(0) | 0(0) | 0(0) | 0(0) | 0(0) | 0(0) |
| Reduced Vision | 12 (6.1%) | 12 (5.9%) | 0(0) | 0(0) | 0(0) | 0(0) | 0(0) | 0(0) | 0(0) | 0(0) |
| Hearing loss | 0(0) | 0 (0) | 3 (0.6%) | 4 (0.6%) | 0(0) | 0(0) | 0(0) | 0(0) | 0(0) | 0(0) |

**Table 3b:** Lesions of chest, back, abdomen, buttocks/groin/genitalia.

| Region | Chest |  | Back |  | Abdomen |  | Buttocks/Groin/Genitalia |  |
| --- | --- | --- | --- | --- | --- | --- | --- | --- |
| What Problem he/she had | Children | Lesions | Children | Lesions | Children | Lesions | Children | Lesions |
| Wound injury related | 6 (6.0%) | 6 (5.9%) | 2 (2.6%) | 2 (2.3%) | 2 (0.5%) | 2 (0.4%) | 4 (2.5%) | 4 (2.4%) |
| Wound not injury related | 20 (20.0%) | 21 (20.6%) | 23 (29.5%) | 25 (28.7%) | 14 (3.2%) | 16 (3.2%) | 43 (26.7%) | 46 (27.2%) |
| Burn | 4 (4.0%) | 4 (3.9%) | 7 (9.0%) | 7 (8.0%) | 7 (1.6%) | 7 (1.4%) | 13 (8.1%) | 13 (7.7%) |
| Mass or growth (solid) | 23 (23.0%) | 24 (23.5%) | 37 (47.4%) | 44 (50.6%) | 10 (2.3%) | 10 (2.0%) | 32 (19.9%) | 36 (21.3%) |
| Mass or growth (reducible) | 0(0) | 0(0) | 0(0) | 0(0) | 16 (3.7%) | 16 (3.2%) | 14 (8.7%) | 14 (8.3%) |
| Deformity congenital | 13 (13.0%) | 13 (12.7%) | 5 (6.4%) | 5 (5.7%) | 14 (3.2%) | 15 (3.0%) | 16 (9.9%) | 16 (9.5%) |
| Deformity acquired | 8 (8.0%) | 8 (7.8%) | 4 (5.1%) | 4 (4.6%) | 4 (0.9%) | 4 (0.8%) | 4 (2.5%) | 4 (2.4%) |
| Foreign body | 13 (13.0%) | 13 (12.7%) | 0(0) | 0(0) | 0(0) | 0(0) | 0(0) | 0(0) |
| Cardiac (congenital) | 13 (13.0%) | 13 (12.7%) | 0(0) | 0(0) | 3 (0.7%) | 3 (0.6%) | 0(0) | 0(0) |
| Abdominal distention or pain | 0(0) | 0(0) | 0(0) | 0(0) | 105 (24.0%) | 116 (23.5%) | 0(0) | 0(0) |
| Inability to urinate | 0(0) | 0(0) | 0(0) | 0(0) | 202 (46.2%) | 236 (47.9%) | 0(0) | 0(0) |
| Renal stone | 0(0) | 0(0) | 0(0) | 0(0) | 20 (4.6%) | 20 (4.1%) | 0(0) | 0(0) |
| Bleeding (per rectum) | 0(0) | 0(0) | 0(0) | 0(0) | 36 (8.2%) | 41 (8.3%) | 0(0) | 0(0) |
| Bleeding (per penis) | 0(0) | 0 (0) | 0(0) | 0(0) | 4 (0.9%) | 7 (1.4%) | 16 (9.9%) | 17 (10.1%) |

**Table 3c:**
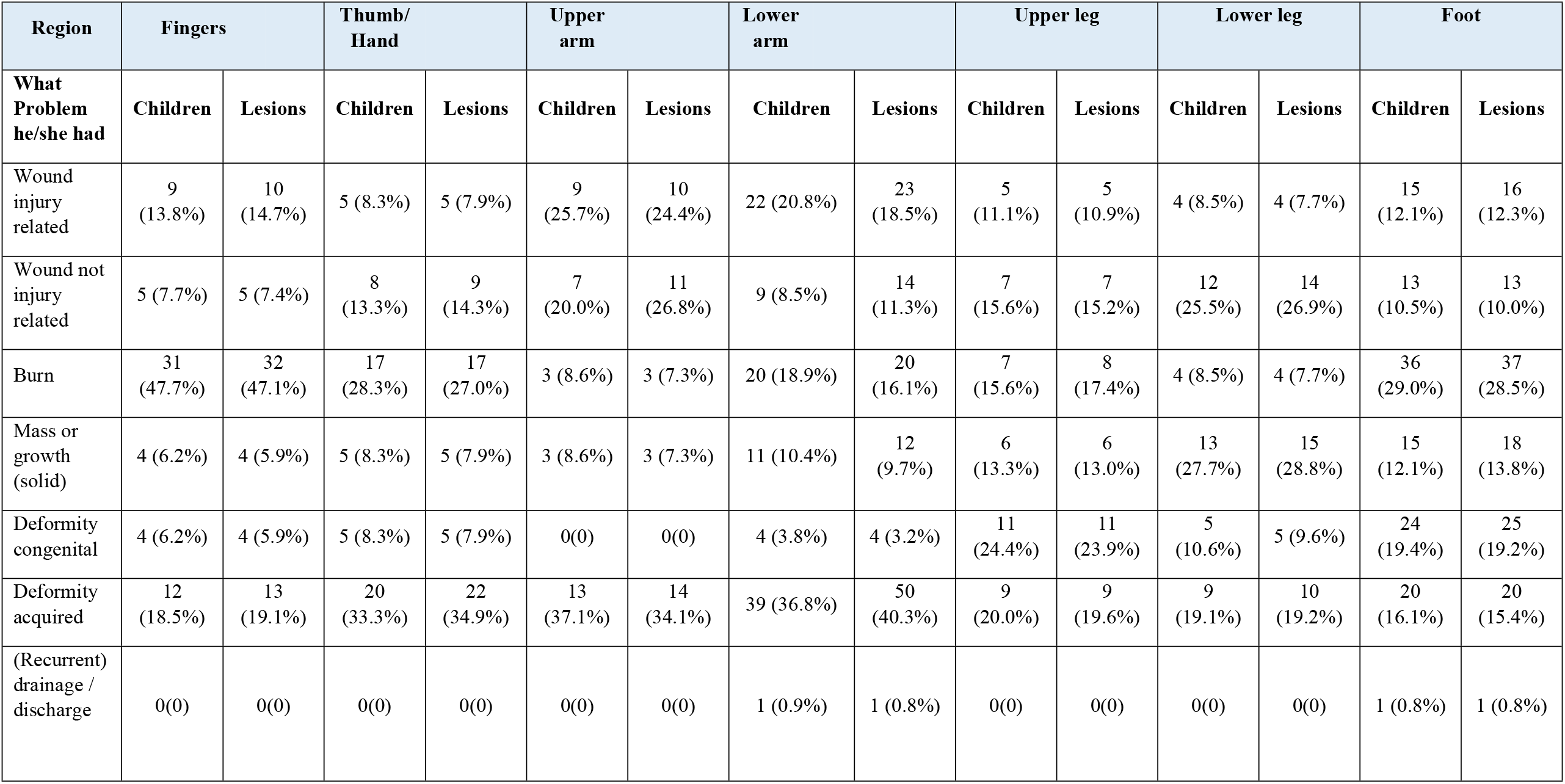
Lesions on Extremities.

| Region | Fingers |  | Thumb/<br>Hand |  | Upper<br>arm |  | Lower<br>arm |  | Upper leg |  | Lower leg |  | Foot |  |
| --- | --- | --- | --- | --- | --- | --- | --- | --- | --- | --- | --- | --- | --- | --- |
| What<br>Problem<br>he/she had | Children | Lesions | Children | Lesions | Children | Lesions | Children | Lesions | Children | Lesions | Children | Lesions | Children | Lesions |
| Wound<br>injury<br>related | 9<br>(13.8%) | 10<br>(14.7%) | 5 (8.3%) | 5 (7.9%) | 9<br>(25.7%) | 10<br>(24.4%) | 22 (20.8%) | 23<br>(18.5%) | 5<br>(11.1%) | 5<br>(10.9%) | 4 (8.5%) | 4 (7.7%) | 15<br>(12.1%) | 16<br>(12.3%) |
| Wound not<br>injury<br>related | 5 (7.7%) | 5 (7.4%) | 8<br>(13.3%) | 9<br>(14.3%) | 7<br>(20.0%) | 11<br>(26.8%) | 9 (8.5%) | 14<br>(11.3%) | 7<br>(15.6%) | 7<br>(15.2%) | 12<br>(25.5%) | 14<br>(26.9%) | 13<br>(10.5%) | 13<br>(10.0%) |
| Burn | 31<br>(47.7%) | 32<br>(47.1%) | 17<br>(28.3%) | 17<br>(27.0%) | 3 (8.6%) | 3 (7.3%) | 20 (18.9%) | 20<br>(16.1%) | 7<br>(15.6%) | 8<br>(17.4%) | 4 (8.5%) | 4 (7.7%) | 36<br>(29.0%) | 37<br>(28.5%) |
| Mass or<br>growth<br>(solid) | 4 (6.2%) | 4 (5.9%) | 5 (8.3%) | 5 (7.9%) | 3 (8.6%) | 3 (7.3%) | 11 (10.4%) | 12<br>(9.7%) | 6<br>(13.3%) | 6<br>(13.0%) | 13<br>(27.7%) | 15<br>(28.8%) | 15<br>(12.1%) | 18<br>(13.8%) |
| Deformity<br>congenital | 4 (6.2%) | 4 (5.9%) | 5 (8.3%) | 5 (7.9%) | 0(0) | 0(0) | 4 (3.8%) | 4 (3.2%) | 11<br>(24.4%) | 11<br>(23.9%) | 5<br>(10.6%) | 5 (9.6%) | 24<br>(19.4%) | 25<br>(19.2%) |
| Deformity<br>acquired | 12<br>(18.5%) | 13<br>(19.1%) | 20<br>(33.3%) | 22<br>(34.9%) | 13<br>(37.1%) | 14<br>(34.1%) | 39 (36.8%) | 50<br>(40.3%) | 9<br>(20.0%) | 9<br>(19.6%) | 9<br>(19.1%) | 10<br>(19.2%) | 20<br>(16.1%) | 20<br>(15.4%) |
| (Recurrent)<br>drainage /<br>discharge | 0(0) | 0(0) | 0(0) | 0(0) | 0(0) | 0(0) | 1 (0.9%) | 1 (0.8%) | 0(0) | 0(0) | 0(0) | 0(0) | 1 (0.8%) | 1 (0.8%) |

#### Chest

In the 102 lesions identified on the chest, 24 (23.5%) comprised of a mass or growth, closely followed by 21 (20.6%) non-injury related wounds. Most lesions were not secondary to an accident (n = 83, 81.4%) but amongst the remaining 19 (18.6%) that occurred following an accident, falls were identified as one of the main reasons (n = 10, 52.6%). The unmet surgical need of lesions on the chest was found to be the lowest, amounting to only 5.9%.

#### Back

We identified 87 lesions on the back, of which approximately half consisted of mass or growth (n = 44, 50.6%). Only 12 (13.8%) lesions were reported to have started after an injury/accident, while the remaining 75 (86.2%) lesions were not secondary to an accident. In 12 injury related wounds, burns from hot liquid or object were seen as the most common cause (n = 5, 41.7%). The unmet surgical needs for the lesions on the back was found to be 6.9%.

#### Abdomen

In the 493 abdominal problems noted, the majority consisted of a child unable to urinate (236, 47.9%) followed by abdominal distention or pain (n = 116, 34.1%). A total of 481 (97.6%) lesions were not preceded by an accident. Wounds secondary to an accident mostly resulted from burns with hot objects (n = 5, 41.7%). The unmet surgical need for the abdominal lesions was 13.4%.

#### Buttocks/ Groin/ Genitalia

In a total of 169 lesions identified, the majority consisted of wounds that did not result from an injury (n = 46, 27.2%) followed by a solid mass or growth (n = 36, 21.3%). A sum of 19 lesions resulted from an accident or injury, with hot object burns (n = 8, 42.1) accounting for most, followed by fall (n = 4), animal attack (n = 3) or open fire explosion (n = 3). One child reported a stab/slash/crush related injury. The unmet surgical need for the lesions on the buttocks/groin/genitalia was found to be 14.8%. Details of lesions of chest, back, abdomen and buttock / groin / genitalia are available in **Table 3b**.

#### Extremities

In the 296 lesions on the upper limb, proximally to distally 13.9% were on the upper arm, 41.9% on the lower arm, 21.3% on the thumb and hands and 23% were on the fingers. Majority lesions of the upper arm (34.1%), lower arm (40.3%) and thumb (34.9%) were due to acquired deformities, while fingers were most affected by burns (47.1%). The upper limb lesions were mostly due to an injury or accident (n = 204, 68.9%). Of the lesions secondary to injury, falls were the primary cause in the upper arm, lower arm, and hand/thumb, whereas for fingers hot object related burns accounted for the highest percentage of the total lesions.

Overall, 228 (9.3%) lesions were found on lower limbs. Proximally to distally, lesions were distributed as follows; upper leg (n = 46, 20.2%), lower leg (n = 52, 22.8%) and feet (n = 130, 57%). Lesions of the upper legs were mainly deformities, both congenital (n = 11) and acquired (n = 9), while the lower legs were mostly affected by abnormal growths (n = 15, 28.8%), and burns were the most frequently occurring lesions of the feet (n = 37, 28.7%).

The total unmet surgical need for the combined upper and lower limb lesions was found to be 11.8%. Further details about lesion on extremities is shown in **Table 3C**.

#### Photographs

A sum of 233 photographs were taken of children with lesions. These pictures were shared with surgeons at the Aga Khan University Hospital for scoring and obtaining preliminary diagnoses via interdepartmental collaboration of multiple surgery faculty members. Majority of the pictures were of children with lesions on the head and neck region, followed by extremities and least from children with lesions on the back. More information about the diagnosis can be found in **Appendix 1**.

### Health Seeking Behavior

A large percentage of the lesions (n = 2,632, 85.7%) were managed at a health care facility, ranging from private (52.4%), government (33.8%) and NGO based setup (0.2%), while the remaining 7.8% lesions were managed at home, or by a Hakeem/traditional healer (5.9%). Treatment essentially consisted of only medical management (n = 2,289, 87%). Surgical treatment was only received for 291 (11%) lesions, which included minor procedures, major procedures, traction and manipulation /casting /sling. Fifty-two (2%) lesions did not receive any care. **Table 4** shows a more detailed picture of the health seeking behavior.

**Table 4:**
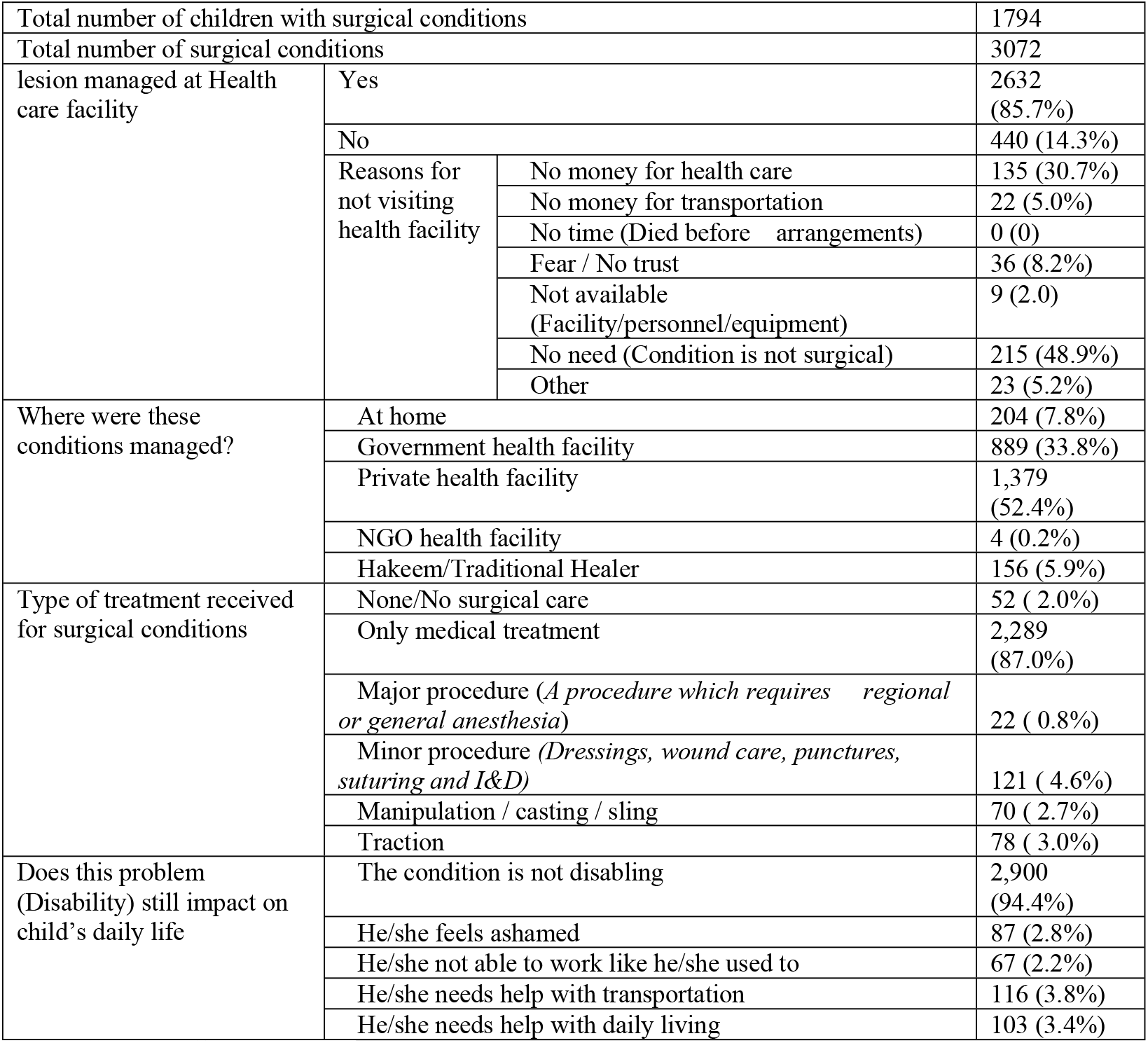
Lesion management and impact.

The perception of the condition being non-surgical (n = 215, 48.9%) and an inability to afford health care (n = 135, 30.7%) were found to be the major reasons for parents not taking their children to a health facility. The data depicts that a vast majority of the conditions reported were not disabling for the children (n = 2,900, 88.6%). A few children reported needing help with transportation (3.5%), needing help with daily living (3.1%), feeling ashamed of their condition (2.6%), and not being able to work the way they were previously able to (2%).

### Health Facility Assessment

A total of 39 health facilities were identified and surveyed in the TMK district. They consisted of one District Headquarter Hospital (DHQ), three Rural Health Clinics (RHC), 14 Basic Health Units (BHU) and 21 District Council Dispensaries (DCD). These facilities were assessed using Pediatrics Personnel, Infrastructure, Procedures, Equipment and Supplies (Pedi PIPES) survey form.

#### Personnel

Trained staff was only identified to be present at the DHQ; the staff present comprised of three anesthesiologists, two pediatricians with two pediatric trained nurses and one general surgeon. There were no nurse anesthetists. No pediatric surgeon or a medical doctor able to operate on children was available.

#### Infrastructure

The facilities comprised of 197 beds of which 22 were allocated for pediatric patients. All 39 facilities had an electricity supply (external source or generator). Other infrastructure components that were present in most of the facilities included running water and medical records. Laboratories to test blood and urine were available in 89.7% of the facilities surveyed. A very small proportion of the hospitals were found to have incinerators (23.1%), ultrasonography (20.5%), plain radiography (12.8%), post-operative care area (12.8%), an emergency department (10.3%) and special care baby unit (10.3%). Less than 10% of the hospitals had operating rooms (7.7%), blood banks (2.6%) and pediatric ventilators (2.6%). None of the facilities had a neonatal or general intensive care unit (ICU), a newborn incubator, or a computed tomography (CT) scan machine. Details have been mentioned in **Table 5**.

**Table 5:** Infrastructure items.

| Infrastructure items | No. (%) * |
| --- | --- |
| Operating room | 3 (7.7) |
| Laboratory (blood and urine) | 35 (89.7) |
| Medical records | 38 (97.4) |
| Emergency department | 4 (10.3) |
| Newborn incubator | 0 (0) |
| Postoperative care area | 5 (12.8) |
| Blood bank | 1 (2.6) |
| Plain radiography | 5 (12.8) |
| Ultrasonography | 8 (20.5) |
| Special Care Baby Unit | 4 (10.3) |
| Electricity (external source or generator) | 39 (100) |
| Incinerator | 9 (23.1) |
| Running water | 38 (97.4) |
| NICU or General ICU | 0 (0) |
| Pediatric ventilator | 1 (2.6) |
| Computed tomography | 0 (0) |

#### Procedures

At the time of the survey, 14 different types of procedures were being performed at the health facilities. Basic and less intricate procedures were performed more than the complex procedures. Suturing (94.9%), wound debridement (94.9%) and incision and drainage of abscess (82.1%) were frequently performed. Less commonly performed procedures included resuscitation (20.5%), burn management (10.3%), traction of closed fracture (2.6%), casting of fracture (2.6%), pediatric hernia repair (2.6%), ovarian cystectomy (2.6%), appendectomy (2.6%) and spinal/ general/ ketamine induced anesthesia (2.6%) each. Details available in **Table 6**

**Table 6:** Procedures.

| Anesthesia<br>No. (%) | Respiratory<br>No. (%) | Gastrointestinal<br>No. (%) | Urogenital<br>No. (%) | Orthopedic<br>No. (%) | Unclassified<br>No. (%) |
| --- | --- | --- | --- | --- | --- |
| Ketamine<br>1 (2.6) | Chest tube<br>insertion<br>1 (2.5) | Appendectomy<br>1 (2.6) | Pediatric<br>hernia repair<br>1 (2.6) | Traction of<br>closed fracture<br>1 (2.6) | Resuscitation<br>8 (20.5) |
| General<br>1 (2.6) |  |  | Ovarian<br>cystectomy<br>1 (2.6) | Casting of<br>fracture<br>1 (2.6) | Suturing<br>37 (94.9) |
| Spinal<br>1 (2.6) |  |  |  |  | Wound<br>debridement<br>37 (94.9) |
|  |  |  |  |  | I&D of<br>abscess<br>32 (82.1) |
|  |  |  |  |  | Burn<br>management<br>4 (10.3) |

#### Equipment and Supplies

Most hospitals had an appropriate availability of equipment. We found more than 90% of the hospitals had a supply of examination gloves, aprons, adhesive tape, thermometers, stethoscopes, infant weighing scale, syringes, intravenous cannulas, oxygen mask and tubing, compressed oxygen, tourniquet, sterile gloves/gauze, facemask, pulse oximeter and gowns for surgeon/scrub nurse. Other items that were present in more than half of the facilities included kidney-dishes, sterilizers, disposable needles, theater shoes, absorbable and non-absorbable sutures, oxygen concentrators, 12 F or smaller nasogastric tubes, pediatric blood pressure cuff, intravenous fluid infusion set and manual or electric suction pumps. Apnea monitors were not available at any of the hospitals. The availability of anesthesia machines, operating room lights, electrocautery machine, scalpel blades, syringe pumps, and sterile drapes was notably low, while laparoscopic surgery supplies, chest tubes, neonatal T-piece and endoscopes were unavailable in all 39 facilities. Details available in **Table 7** and **Table 8**

**Table 7:** Equipment’s.

| **Equipment** | **No. (%)** |
| --- | --- |
| Thermometer | 38 (97.4) |
| Stethoscope | 38 (97.4) |
| Endotracheal tube (pediatric) | 5 (12.8) |
| Weighing scale (infant) | 38 (97.4) |
| Oropharyngeal airway (pediatric) | 7 (17.9) |
| Anesthesia machine | 2 (5.1) |
| Kidney dish | 35 (89.7) |
| Sterilizer (autoclave) | 32 (82.1) |
| Oxygen mask and tubing | 38 (97.4) |
| Suction pump (manual or electric) | 24 (61.5) |
| Operating room light | 4 (10.3) |
| Surgical instrument set (abdominal) | 5 (12.8) |
| Electrocautery machine | 1 (2.6) |
| Compressed oxygen in cylinder | 38 (97.4) |
| Bag-valve mask (pediatric) | 19 (48.7) |
| Pulse oximeter | 37 (94.9) |
| Oxygen concentrator | 31 (79.4) |
| Blood pressure measuring equipment (pediatric cuff) | 20 (51.3) |
| Neonatal T-piece | 0 (0) |
| Endoscope (esophagoscope/ bronchoscope/ cystoscope) | 0 (0) |
| Syringe pumps | 2 (5.1) |
| Apnea alarm detector/apnea monitor | 0 (0) |

**Table 8:** Supplies.

| Supplies | No. (%) |
| --- | --- |
| Gloves (sterile) | 36 (92.3) |
| Gloves (examination) | 39 (100) |
| Nasogastric tubes 12F or smaller | 20 (51.3) |
| Intravenous fluid infusion sets | 24 (61.5) |
| Blood transfusion sets | 10 (25.6) |
| IV cannulas | 38 (97.4) |
| Syringes | 38 (97.4) |
| Disposable needles | 32 (82.1) |
| Tourniquet | 38 (97.4) |
| Sterile gauze | 37 (94.9) |
| Bandages sterile | 37 (94.9) |
| Adhesive Tape | 39 (100) |
| Suture (absorbable) | 30 (76.9) |
| Suture (non-absorbable) | 34 (87.2) |
| Urinary catheters (must include 6F) | 8 (20.5) |
| Sharps disposal container | 4 (10.3) |
| Face masks | 37 (94.9) |
| Eye protection (goggles, safety glasses) | 11 (28.2) |
| Apron | 39 (100) |
| Boots (theatre shoes) | 34 (87.2) |
| Gowns (for surgeon/scrub nurse) | 36 (92.3) |
| Drapes (for operations) | 1 (2.6) |

#### PediPIPES scores and indices

PediPIPES personnel, infrastructure, procedures, equipment, and supplies score were calculated. The hospital with the highest PediPIPES indices in Tando Mohammad Khan had the following: Personnel score 8, infrastructure score 9, procedure score 12, equipment score 17 and supplies score of 21.

## DISCUSSION

SOSAS was primarily developed to measure surgical disease burden in low- and middle-income countries (10). From the time of its creation and first use, SOSAS has become a validated tool to assess a vast range of surgically treatable conditions at the population level (10, 11, 14). This execution of the SOSAS and PediPIPES survey in the rural district of Tando Mohammad Khan is the first of its nature in Pakistan. The primary aim of this study was to lay out an introductory population-based data on the burden of surgical disease in the rural areas of Pakistan, which will contribute to our comprehension of the epidemiology of surgical diseases in LMICs.

Of the reported child death in the last five years in Tando Mohammad Khan, 36.3% of the deaths were due to lesions that required surgical attention. The majority of children (approximately 80%) were taken to a health care facility (private more frequently that public facilities), but only 3.4% children received surgical care. This identifies a high unmet surgical need in the region. A large proportion of children with surgical lesions were taken to a health care facility for management, but only a small fraction (11%) were surgically managed, while the majority (87%) received only medical management. On assessment of why children with surgical lesions were not taken to a health facility, it was found that a high proportion of the population did not perceive the condition as requiring surgical management. The next most frequent reason for not seeking care was the inability to afford health care.

Health facility assessment done through PediPIPES identified that only one facility in Tando Mohammad Khan had trained staff, comprising of anesthesiologists, pediatricians, nurses and one general surgeon. This identifies a lack of skilled personnel available to cater to the surgical need of children in the area. Amongst the total 39 facilities in the area, only 22 paediatric beds were available. These facilities had the capacity to perform only basic procedures such as suturing, wound debridement and incision and drainage.

Until recently, global health attention was mainly focused on addressing communicable diseases and maternal and child mortality in LIC and LMIC (5). In contrast, this study highlights the significant surgical burden that exists and the importance of developing a health care structure to manage it. Compared to other LMICs (11, 12, 14, 15, 26)the rates of current and unmet surgical need in Tando Mohammad Khan are high. Pakistan currently faces a challenge of inadequate surgical care delivery to children (27). According to the World Bank 2019 data, within South Asia, India was reported to have a lower number of physicians per 1000 individuals compared to Pakistan (0.7 vs. 1.1) (28). However, in correlation to the SOSAS study conducted in 2019 by Cherukupalli et al. (5), the unmet surgical need in rural India was reported to be 6.5%, which is much lower than a staggering 14.3% in Pakistan as identified in this study.

Access to surgical care could be addressed with the continuing development of surgical capacity at lower-level facilities, improving referral systems, and surgical training of non-physicians. Malawi, Mozambique and other countries have invested in training of non-physicians in surgery. However only a small fraction of the need can be dealt with in this fashion (29). In addition to health facility capacity development, community-based educational programs for strengthening pre-hospital systems are vital (5). The implementation of first responder courses (30) in Tando Mohammad Khan could train lay-people to perform initial stabilization and transportation of the injured to a higher level of care. Such programs have been shown to reduce mortality and physiological severity scores in trauma patients, and can improve pre-hospital infrastructure by empowering community members with basic emergency and trauma care skills (30). Health service delivery may be greatly improved through a simple reorganization of services without any cost. For example, this can be demonstrated by the essential trauma-care guidelines and their use as needs assessment tools in multiple countries (31). Increasing the availability of medications, equipment, supplies, and banked blood at the primary and secondary health care centers are essential steps as well (4, 32). This allows adequate care for basic surgical problems at lower-level hospitals, whilst also providing an opportunity to stabilize more critically ill patients prior to being transported to a tertiary care center for more complex procedures.

This study has some potential limitations. One significant limitation is in-built in the survey’s definition of operative conditions. All operative conditions in this survey relied upon participants’ self-reported conditions in the verbal interview. Thus, the conditions identified through this survey serve as proxies in estimating operative disease prevalence. This may overestimate some conditions, which may require follow up rather than surgical intervention. This survey also does not determine conditions such as cancers, which could require surgical care, thus underestimating the true surgical burden of disease. Despite these limitations, surgical conditions recognized by SOSAS provide an initial estimate of need for surgical consultations.

After the success of an initial survey in rural district of Pakistan, the next step will be to perform a larger countrywide survey in Pakistan with an aim to compare the burden of surgical conditions in urban and rural regions. We also need to explore the differences in community’s approach to health seeking and the treatment options available. The successful implementation of photographs taken on-site and using them to determine a diagnosis with the help of physicians from different sub-specialties can be followed up by incorporation of a physical exam performed by a local physician for further validation in future studies. The incorporation of a physical exam would help confirm verbal interview findings and strengthen the SOSAS tool.

## Data Availability

All data generated or analysed during this study are included in this published article and its supplementary information file.

